# Oral and gut microbiome biomarkers of susceptibility to respiratory tract infection in adults: a longitudinal cohort feasibility study

**DOI:** 10.1101/2021.12.21.21268189

**Authors:** Claire A. Woodall, Ashley Hammond, David Cleary, Andrew Preston, Peter Muir, Ben Pascoe, Samuel K. Sheppard, Alastair D. Hay

## Abstract

**Background and aim:** Respiratory tract infections (RTIs) are common in the community. There is some evidence that microbial biomarkers can be used to identify individuals most susceptible to RTI acquisition. We investigated the feasibility of recruiting healthy adults to collect at-home self-reported socio-demographic data and biological samples, saliva (oral) and stool (gut) at three time points (TPs): baseline/start of the study (TP-A), during an RTI (TP-B) and end of study (TP-C).

**Methods:** Healthy adults were recruited from two urban Bristol GP practices. To identify respiratory pathogens in all saliva samples and RTI-S stool samples reverse transcriptase PCR (RT-PCR) was applied. We compared oral and gut samples from participants who developed RTI symptoms (RTI-S) and those who remained healthy (no-RTI) using 16S rRNA profiling microbiome analysis to identify the core microbiome, alpha and beta diversity, and biomarkers for susceptibility to RTIs from baseline samples (TP-A) when all participants were healthy.

**Results:** We recruited 56 participants but due to the UK COVID-19 pandemic disruption we did not receive samples from 16 participants leaving 19 RTI-S and 21 no-RTI participants with socio-demographic and microbiome data. RT-PCR revealed coagulase-negative *Staphylococcus* carriage was significantly higher in RTI-S participants compared to those who remained healthy and RTI symptoms may have been due to viral influenzae and bacterial co-infection with *Haemophilus influenzae*. Core microbiomes of no-RTI participants contained a greater number of taxa compared to RTI-S participants. Microbial biomarkers of RTI susceptibility in the oral cavity were an increased abundance of the pathobiont *Streptococcus sobrinus* and decreased probiotic bacterium *Lactobacillus salivarius* whereas in the gut there was an increased abundance of the genus *Veillonella* and decreased abundance of *Coprobacillus*.

**Conclusion:** In our feasibility study we found oral and gut microbial biomarkers for susceptibility to RTI acquisition. Strategies to identify those most vulnerable to RTI in the community could lead to novel interventions to decrease respiratory infection and associated health services burden.

## Introduction

Respiratory tract infections (RTIs) are the most common infections seen in primary care, and the single greatest contributor to the overall burden of disease worldwide [1]. In the UK, respiratory illness costs the National Health Service (NHS) £11 billion per year [2]. Seasonal community-acquired viral RTIs such as common colds, bronchitis, and influenza-like illness (ILI) are common in adults, who may experience multiple infections per year. In addition, COVID-19 disease caused by severe acute respiratory syndrome coronavirus-2 (SARS-CoV-2) which, now may also be classified as a seasonal RTI and has resulted in an increase burden on frontline NHS services amounting to £4-5bn a year [3]. One way to reduce the burden of RTIs on global health services is to understand which members of the community are most at risk of RTI acquisition so that protective measures can be implemented in a timely manner.

The human microbiome is an important moderator of health and disease. At mucosal surfaces of the gastrointestinal (gut) and respiratory tract a complex bi-directional microbial network influences the host immune-regulatory mechanisms and homeostasis [4]. Microbial signature patterns at this ‘gut-lung’ axis are increasingly reported as having an association with disease susceptibility [5]. Growing evidence suggests microbial biomarkers of susceptibility to RTI are identifiable, yet many more studies in this area are needed [6]. A holistic insight into the role of microbes before, during and after an RTI offers enormous potential for clinical diagnostic interventions and greater insight into infection prevention.

Here, we aimed to conduct a feasibility study to identify microbiome biomarkers of susceptibility to community-acquired RTI in healthy adults. We investigated longitudinal patterns of gut and oral microbiomes from stool and saliva samples respectively, collected from adult participants who acquired an RTI (RTI-S) during the study compared to those who remained healthy (no-RTI). Our findings demonstrate the feasibility of recruiting and retaining urban community participants, who performed self-collection of samples and self-reported RTI symptoms. Microbiome analysis revealed putative biomarkers of RTI susceptibility particularly within the respiratory tract niche.

## Methods

### Ethical declaration

The study was approved by the Southwest – Central Bristol Research Ethics Committee (REC Reference 19/SW/0167) on 2^nd^ October 2019. NHS Health Research Authority approval was granted on 22^nd^ October 2019.

### Participant recruitment

Primary care practices were invited to express interest in taking part in the study via the National Institute for Health Research (NIHR) Clinical Research Network (CRN). Practices were asked to send out study invitations to healthy patients aged between 18 and 70 years and living independently, using diagnostic codes in the medical records to exclude patients that present conditions which are considered confounders for the gut and oral microbiome, such as: a condition affecting the immune system including but limited to, lupus; without capacity to consent; with a severe life-limiting illness; and those with conditions known to predispose to infection (chronic lung disease, cystic fibrosis, and type 1 and 2 diabetes and who are at greater risk of developing community-acquired RTIs including, but not limited to, chronic obstructive pulmonary disease [COPD], asthma, and bronchiectasis). The practices mailed eligible patients an invitation letter, consent form, and participant information sheet. Those who were willing to participate were asked to return their signed consent form to the study team using prepaid envelopes. Participant recruitment took place between November 2019 and January 2020. The study was closed prematurely in May 2020 due to the onset of the COVID-19 pandemic in the UK.

### Metadata collection

Upon receipt of the participant signed consent form, the study team emailed or telephoned the patient to confirm eligibility and provide instructions regarding baseline data completion online, after which point the participant was fully enrolled. Participants were requested to complete a questionnaire to collect covariates used as metadata for microbiome analysis: age, sex, height, weight, RTI symptoms, ethnicity, education, employment, public transport usage, history of RTIs in previous 12 months, handwashing frequency, smoking, consumption and frequency of alcohol, meat, fruit, vegetables, probiotics, herbal and/or homeopath supplements, over-the-counter medication, number and age of adults and or children in the household, how many children in day-care and/or pre-school, smoker in the home, pet type and ownership, RTI symptoms, GP visits, antibiotics taken for this RTI.

Self-reported RTI symptoms were entered into the database in response to weekly e-mails requesting participants to respond with a ‘yes’ or ‘no’ to the query asking if they had developed at least one or more ‘new’ RTI symptom at any time during the study. Symptoms included: a blocked and/or runny nose, earache and/or ear discharge, sore throat, cough, chest symptoms including breathing faster than normal, wheeze and/or whistling chest. A positive response prompted the participant to complete a short symptom resolution questionnaire, which included information on what symptoms they had and when they started, whether they consulted with their primary care clinician for their symptoms, took any medication for their symptoms, and took time off work due to their symptoms. From this point, weekly emails commenced asking participants whether their symptoms had fully resolved. Once symptoms had fully resolved the participant active study period ended. A negative response resulted in no further action and weekly emails continued asking whether they had developed new RTI symptoms for the duration of the study, or until they responded ‘yes’. For participants who did not report any RTI symptoms and remained healthy during the study, they continued to receive weekly symptom emails until the end of the study period.

At the end of the study, all participants including those who reported RTI symptoms (RTI-S) and those that did not (no-RTI) were prompted to complete a brief end of study questionnaire. This was a condensed version of the symptom resolution questionnaire which simply asked the participant for feedback on the study. Once completed, the participant active study period ended.

### Stool and saliva collection

Stool and saliva samples were self-collected by each participant at three time points (TP) and posted directly to the study team:

- TP-A: Baseline samples. Collected within 3 days of recruitment to the study if the participant was RTI symptom-free. If the participant had RTI symptoms at recruitment, they were asked to collect their samples once they had been free of symptoms for at least two days.
- TP-B: Symptomatic samples. Collected at onset of RTI symptoms or within 2 days of RTI symptoms being reported and reminders were emailed to participants to collect their samples if they reported having RTI symptoms.
- TP-C: Recovery/post-RTI for participants who acquired an RTI and were collected once the participant reported symptoms had resolved fully for at least 2 consecutive days. Or end of study samples for participants who remained healthy.

Stool and saliva samples were collected into OMNIgene.GUT and OMNIgene.ORAL (DNAGenotek) collection tubes, respectively. Participants were provided with detailed sample collection instructions, including a link to the manufacturer’s video instructions. Specimens were packaged into boxes and posted by Royal Mail directly to the research laboratory for bacterial genomic DNA (gDNA) extraction and subsequent sequencing for microbiome analysis. An aliquot of each saliva and RTI-S stool specimen was transported on ice to the clinical diagnostics laboratory for reverse-transcriptase-PCR (RT-PCR) analysis.

### Microbiome profiling

Bacterial genomic DNA (gDNA) was extracted using QIAamp PowerFecal Pro DNA kit (QIAGEN) according to the manufacturer’s instructions. Bacterial gDNA was quantified using a Qubit™ fluorometer, using the double stranded DNA, Broad Range, reagents according to the manufacturer’s instructions (Life Technologies, Carlsbad, CA). The V4 region of the bacterial 16S rRNA gene was amplified using V4 primers 515F (5’ – GTGCCAGCMGCCGCGGTAA – 3’) and 806R (5’ – GGACTACHVGGGTWTCTAAT - 3’) to generate an amplicon of 300 bp by Novogene Europe. Amplicons were barcoded and sequenced on Illumina NovaSeq 6000, reads were trimmed of barcodes and primers and the resulting pair ended sequence was 250 bp.

### Bioinformatics and statistical analysis

The sequence reads were processed using Quantitative Insights into Microbial Ecology tool (QIIME2-2020.2) [7]. Raw sequences were denoised and chimeras removed with DADA2 [8]. To improve the accuracy of phylogenetic placements, amplicon sequence variants (ASVs) were aligned with mafft [9] (via q2-alignment) and constructed using the fasttree2 [10]. Taxonomic classification was preformed using the q2-feature-classifer [11] classify-sklearn naïve Bayes taxonomy classifier and Greengenes 13_8 (99%) reference sequences [12]. The feature table, taxonomy, phylogenetic tree and sample metadata were then combined into a Phyloseq object using QIIME2R [13] via qza_to_phyloseq.

All further analysis was done in R (v3.6.0) [14] via RStudio (v1.3.1073) using microbiome scripts. Figures were produced using the package ggplot2 [15]. Phyloseq (v1.29.077) [16] was used following a published workflow [17]. Potential sample contaminants were identified by prevalence in negative control samples and removed using the R package ‘decomtam’ [18]. Sample processing, and bioinformatic analyses were tested using samples spiked with *Bordetella sp*. prior to DNA extraction. ASVs with Phylum classifications of ‘NA’ or ‘uncharacterised’ and any Phyla with a total of fewer than five ASVs whose provenance suggested low level contamination were removed using the subset_taxa() function. Alpha diversity was performed using the estimate_richness() and stat_compare_means() was used to compare TP groups using the nonparametric Wilcoxon test. Beta diversity was determined by plotbeta for hierarchical clustering by Bray–Curtis dissimilarity using Principal component analysis (PCoA). Betatest() function was used to calculate the permutation multivariate analysis of variance (PERMANOVA) values for variables of interest. Core microbiome analysis was performed using the ‘microbiome’ vignette using heatmaps visualisation [19]. The Linear Discriminant Analysis (LDA) coupled with the effect size (LEfSe) [20] was used to find taxa that were significantly different (LDA score > 2, *p* <0.05). The ldamarker () function was used for LEfSe analysis based on Kruskal-Wallis and LDA analysis.

### Diagnostics of common respiratory microbes

Upon clinical laboratory receipt of saliva and stool samples, 100µl of each sample was extracted using QIAsymphonySP (QIAGEN, UK) along with internal quality controls containing bacteriophages T4 and MS2 using the QIAsymphony DSP Virus Pathogen Mini Kit (QIAGEN, UK) and 60µl elution protocol. Low volume saliva samples (<200µl) were diluted with 200µl phosphate buffered saline and recovered by low-speed centrifugation into a 2ml collection tubes.

Here, sample collection overlapped with the COVID-19 pandemic in the UK so it was possible samples were infected with the SARS-CoV-2 coronavirus. To determine the presence of coronavirus in all saliva samples and RTI-S stool samples we used RT-PCR for detection of the Beta-CoV E gene and SARS-CoV-2 S gene targets. A clinically validated 42-microbe low density TaqMan Array Card (TAC) (Applied Biosystems, Foster City, CA, USA) was used to detect common respiratory microbes, exogenous internal controls (T4 and MS2 bacteriophage) and endogenous human control genes (18S rRNA and RNase-P) [21]. Samples were run in 3 batches, amplified, and analysed using a Life Technologies Custom TaqMan Low Density Array system on an Applied Biosystems Life Technologies ViiA-7TM real-time PCR system as described elsewhere [22]. A cycle threshold (Ct) value <38 for any gene target was reported as a positive result. Microbe carriage was calculated as the number of positive results as a percentage out of all positive results. The Pearson’s chi-squared test statistic (95% CI) was used to determine significance between observed proportions of microbial carriage.

## Results

### Study population characteristics

Two GP practices were recruited to the study serving a broad range of socio-economic populations, both within central Bristol, England. The study recruitment and data collection flow-chart describes stool and saliva sample collection from each study participant at three TPs (Figure 1).

**Figure 1.**
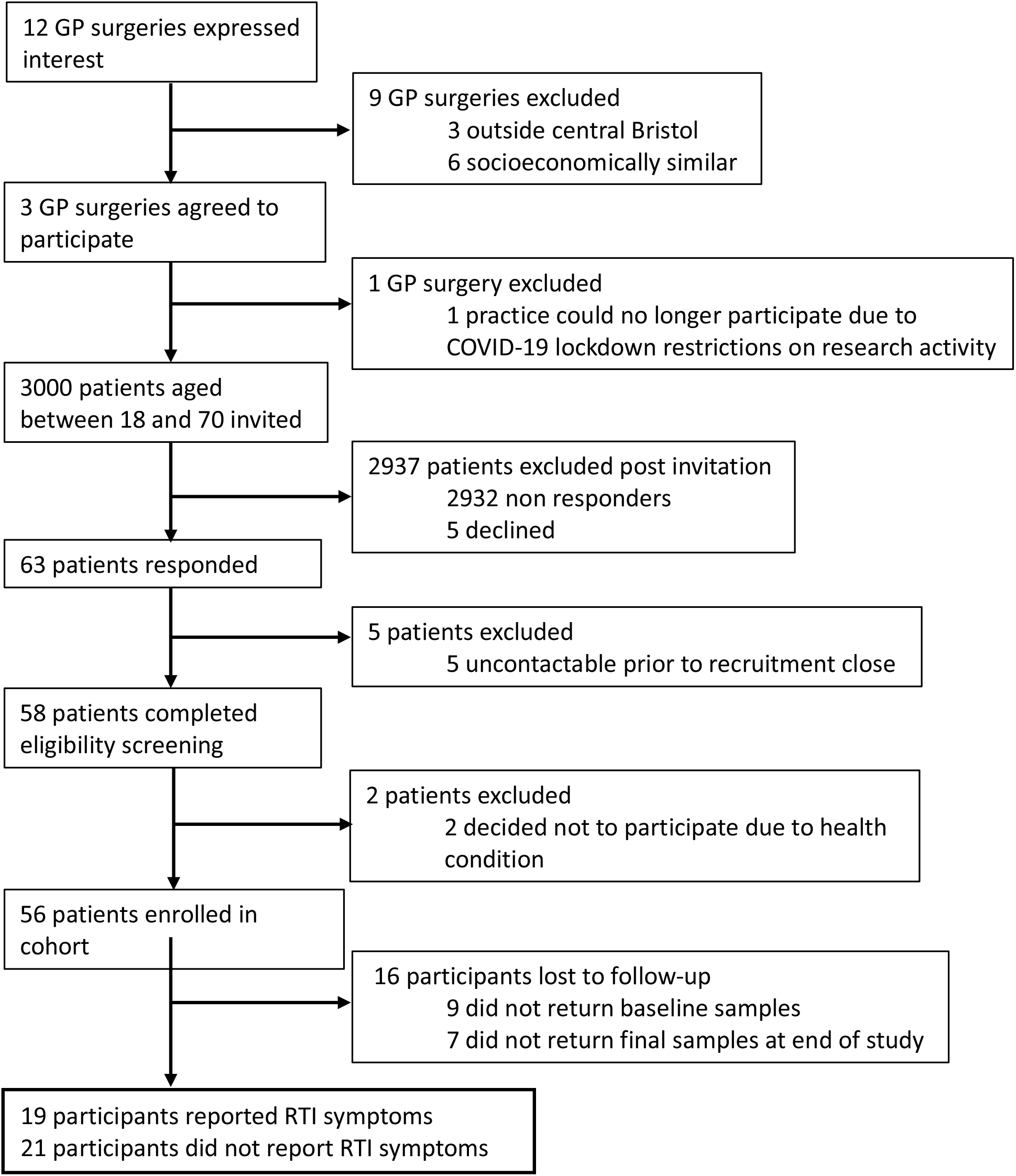
Participant recruitment flowchart

Between October 2019 and March 2020, both stool and saliva samples were collected from 56 participants. A total of 9 participants either did not collect and return TP-B or TP-C samples, or samples were lost in transit because of the UK national Covid-19 lockdown. Therefore, it was not possible to determine whether these 9 participants were RTI-S or no-RTI status. Of the remaining 19 RTI-S participants a further 2 sample sets at TP-C were lost in transit due to postal disruption by the UK COVID-19 pandemic lockdown. We intended to collect health-related risk factors from participant medical notes, including current medications and antibiotic prescriptions in the 12 months prior to recruitment, however, due to the pandemic we were unable to access these data. Descriptive statistics for the most relevant covariates from 40 participants are shown in Table 1.

**Table 1.** Participant characteristics. Stool and saliva samples were collected from participants with RTI symptoms (RTI-S) and no symptoms (no-RTI). Abbreviations: Not applicable (NA). *Missing data, either samples weren’t received in the laboratory or questionnaire data was incomplete

| Descriptive characteristics | Participants |  |
| --- | --- | --- |
|  | RTI-S | No-RTI |
| Number of participants (*n* = 40) | 19 | 21 |
| Gender (%) |  |  |
| Females | 74 | 50 |
| Males | 26 | 50 |
| Age, years (mean ±SD) | 46.1 (3) | 48.3 (2.8) |
| BMI (mean ±SD) | 24.1 (0.9)* | 24.9 (0.8) |
| Ethnicity (%) |  |  |
| White | 100 | 95 |
| Asian | 0 | 5 |
| Education (%) |  |  |
| Higher education | 95 | 57 |
| Further education | 5 | 43 |
| Is there a smoker at home? Yes (%) | 0 | 9.5 |
| Do you have a pet? Yes (%) | 50* | 38 |
| Most common pet. | Dog/s | Cat/s |
| RTI episodes in previous 12 months (mean ±SD) | 1.7 (0.3)* | 1.8 (0.3) |
| **Samples collected at time point (TP):** |  |  |
| TP-A: Baseline (stool & saliva) | 19 & 19 | 21 & 21 |
| TP-B: RTI (stool & saliva) | 19 & 19 | 0 |
| TP-C: End of study (stool & saliva) | 17 & 17* | 21 & 21 |
| Total samples | 110 | 84 |
| Days from TP-A to TP-B (mean ±SE) | 34.8 (7.5) | NA |
| Days from TP-B to TP-C (mean ±SE) | 46.1 (7.7) | NA |
| Days from TP-A to TP-C (mean ±SE) | 80.9 (7.2) | 63.7 (2.9) |
| **RTI symptoms at TP-B:** |  |  |
| Length of episode, days (mean ±SE) | 14 (2.5) | 0 |
| Severity Likert scale :1 = mild to 10 = severe (mean ±SE) | 5.1 (0.5) | 0 |
| Runny nose: Yes (%) | 63.2 | NA |
| Cough: Yes (%) | 36.8 | NA |
| Sore throat: Yes (%) | 68.5 | NA |
| Chest symptoms: Yes (%) | 5.3 | NA |

For no-RTI participants (*n* = 28) 50% were female and their mean age was 48.3 (± 2.8) years. 42 stool and saliva samples were collected at each of TP-A and TP-C from this group. For the RTI-S participants (*n* = 19), 74% were female and the mean age 46.1 (± 3) years. Stool and saliva samples were collected at TP-A (n=38), B (n=38) and C (n= 34). There was a mean of 34.8 (± 7.5) days from the study start at TP-A until participants reported an RTI symptom, at TP-B. There was a mean of 14 (± 2.5) days between collection of TP-B and TP-C (post-RTI symptom) samples. The main RTI symptom was characterised as a sore throat in 68.5% of cases, followed by a runny nose in 63.2% of cases.

### Carriage of respiratory pathogens

To determine carriage of common respiratory tract pathogens in participants, all saliva samples and RTI-S stool samples were analysed by RT-PCR using a 42 pathogen Taqman Array Card (TAC) including coronavirus Beta-CoV E gene and SARS-CoV-2 S gene targets. Pathogen carriage was defined as the percentage of tests for a specific pathogen that were positive (Table 2).

**Table 2.** Carriage of respiratory pathogens in saliva collected from participants with RTI symptoms (RTI-S) and no symptoms (no-RTI) at three time points, pre-, during- and post-RTI. *A significant observed proportional difference, *p* = 0.049. All RTI-S and no-RTI saliva and RTI-S stool samples tested negative for human coronavirus Beta-Cov, E gene and SARS-CoV-2, S gene. Abbreviations: Coagulase-negative staphylococci (CoNS), time point (TP), pre-RTI, during- and post-RTI was TP-A, B, C respectively.

| Respiratory pathogen | Carriage prevalence percent (%) |  |  |  |  |
| --- | --- | --- | --- | --- | --- |
|  | RTI-S |  |  | No-RTI |  |
|  | TP-A<br>(n = 19) | TP-B<br>(n = 19) | TP-C<br>(n = 17) | TP-A<br>(n = 21) | TP-C<br>(n = 21) |
| CoNS | 73.7* | 84.2 | 76.5 | 42.8* | 47.6 |
| *S. aureus* | 0 | 5.3 | 5.9 | 14.2 | 9.5 |
| *F. necrophorum* | 5.3 | 5.3 | 0.00 | 4.8 | 4.8 |
| *H. influenzae* | 26.3 | 31.6 | 29.4 | 19.0 | 14.2 |
| *M. pneumoniae* | 5.3 | 5.3 | 0.0 | 9.5 | 0.0 |
| *S. pneumoniae* | 26.3 | 10.5 | 23.5 | 23.8 | 9.5 |
| *M. catarrhalis* | 10.5 | 5.3 | 17.6 | 14.2 | 9.5 |
| Parechovirus | 0 | 5.3 | 0.00 | 0 | 0 |
| Rhinovirus | 0 | 5.3 | 5.9 | 0 | 0 |
| Rhinovirus-2 | 5.3 | 5.3 | 5.9 | 0 | 0 |
| Influenza A | 0 | 15.7 | 11.8 | 4.7 | 9.5 |

Coronavirus SARS-Cov2 was not detected in any sample. Influenza (subtype A) carriage in RTI-S participants at TP-A, B and C was zero, 15.7% and 11.8% respectively, compared to 4.7% and 9.5% at TP-A and C among no-RTI participants. At all the time points, for both RTI-S and no-RTI participants, there was carriage of *H. influenzae, S. pneumoniae, M. catarrhalis* and all pathogens had high carriage in RTI-S participants TP-B samples. Taken together these observations suggest viral influenzae and bacterial co-infection with *H. influenzae* may have resulted in participant RTI symptoms.

In RTI-S participants CoNS carriage at TP-A, B and C was 73.7%, 84.2% and 76.5%, respectively, whereas in no-RTI participants it was 42.8% at TP-A and 47.6% at TP-C. For TP-A this difference was significant (Pearson’s chi-squared [95% CI], *p* = 0.049). In no-RTI participants *S. aureus* carriage at TP-A and C ranged from 9.5 – 14.2% compared to RTI-S participants which showed no carriage at TP-A, TP-B at 5.3% and TP-C at 5.9%.

### Oral and gut microbiomes

The core microbiome, diversity and biomarker analysis were used to determine differences between the microbiomes at TP-A of participants who acquired a RTI (RTI-S) and those who remained healthy (no-RTI).

Oral and gut microbiomes (OM and GM) of participants were determined by 16S rRNA V4 region profiling where the OM yielded a median of 104,201 reads per sample of which 7692 ASVs were taxonomically assigned, and the GM had a median of 102,627 reads of which 8009 ASVs were assigned. In both microbiomes the relative proportions of phyla, genera and species at TP-A comparing RTI-S and no-RTI participants can be seen in Figure 2 (relative proportions of taxa at each time point comparing RTI-S and no-RTI participants can be found in supplementary Figure S1A, B for OM and GM respectively).

**Figure 2.**
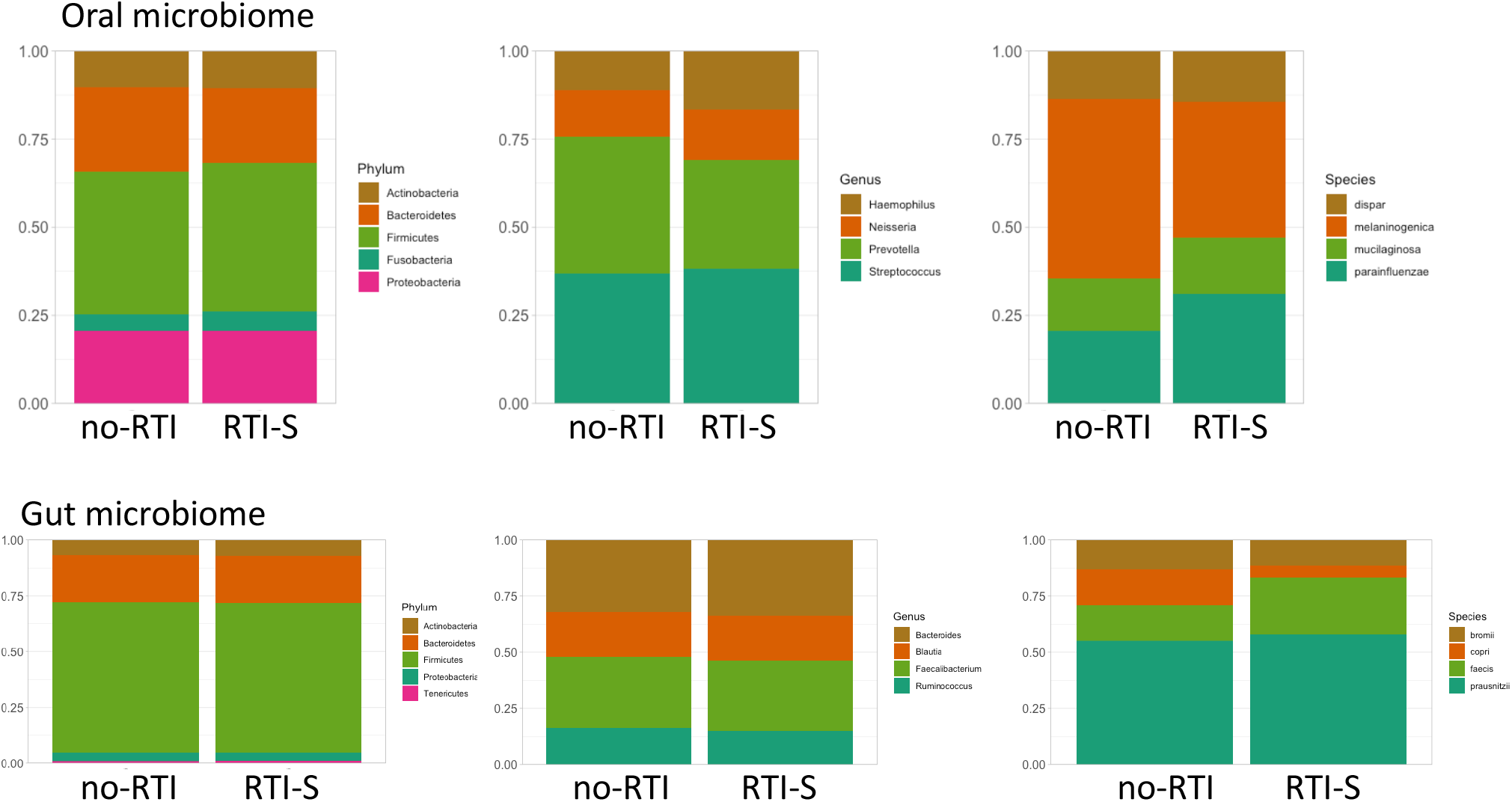
Relative proportions of taxa in oral and gut microbiomes at time point A in participants with RTI symptoms (RTI-S) and no RTI symptoms (no-RTI).

#### Core microbiome

The core taxa of OM and GM (taxa present in 90% of each body niche samples at a relative abundance of at least 1%) were identified (Figure 3). The core OM in RTI-S participants (2,121 total taxa [tt]) was represented by *H. parainfluenzae* (ASV 3988), *Streptococcus* (ASV 5304) and *Neisseria* (ASV 4108) and in no-RTI participants (2846 tt) was *H. parainfluenzae* (ASV 3988), *Streptococcus* (ASV 5304). The core GM in RTI-S participants (3,161 tt) was *Blautia* (ASV 6439) and *F. prausnitzii* (ASVs 6135, 6149) and in the no-RTI participants (3,258 tt) was *Blautia* (ASV 6439).

**Figure 2.**
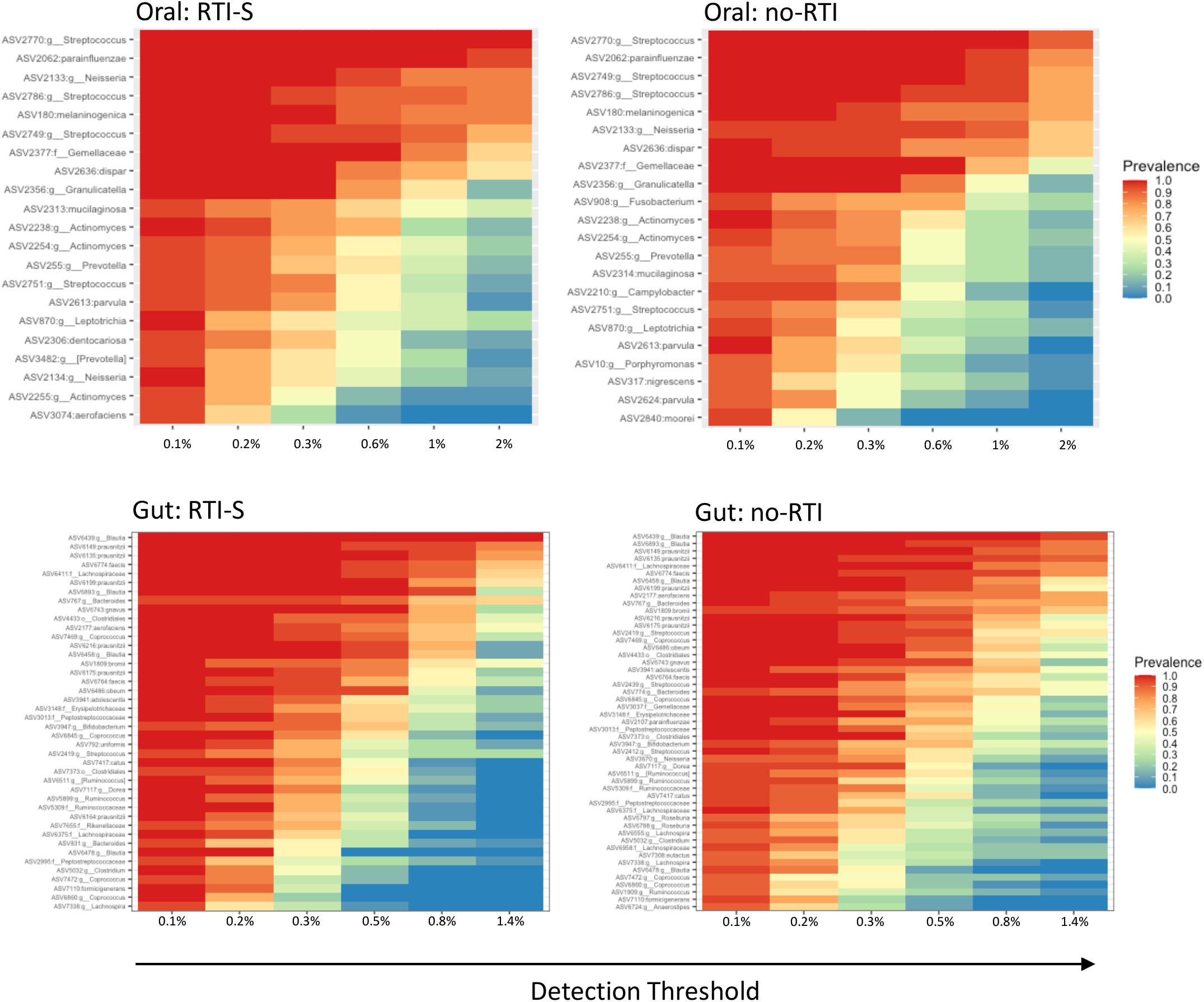
Heatmap representing the core bacterial genera detected across oral and gut microbiomes at time point A in participants with RTI symptoms (RTI-S) and no RTI symptoms (no-RTI).

#### Alpha and beta diversity

Alpha diversity values were measured using the Shannon Diversity Index and Chao1 and significance values for the OM and GM are shown in Table 3 and Figure 4. At TP-A in both microbiomes there were no significant difference in diversity between RTI-S and no-RTI participants. At TP-C in the OM there was a significant increase in diversity between the no-RTI and RTI-S participants and when comparing TP-A and TP-C for only the no-RTI participants. Diversity shifts were not observed for the GM, they remained stable throughout. Interestingly, all saliva samples that tested positive for CoNS (*n* = 62) demonstrated a significant reduction in diversity compared to those that tested negative for CoNS (*n* = 35) (*p* = 2.5 × 10^−3^). Other socio-demographic variables that correlated with a significant difference in diversity are shown in supplementary Table S1.

**Table 3.**
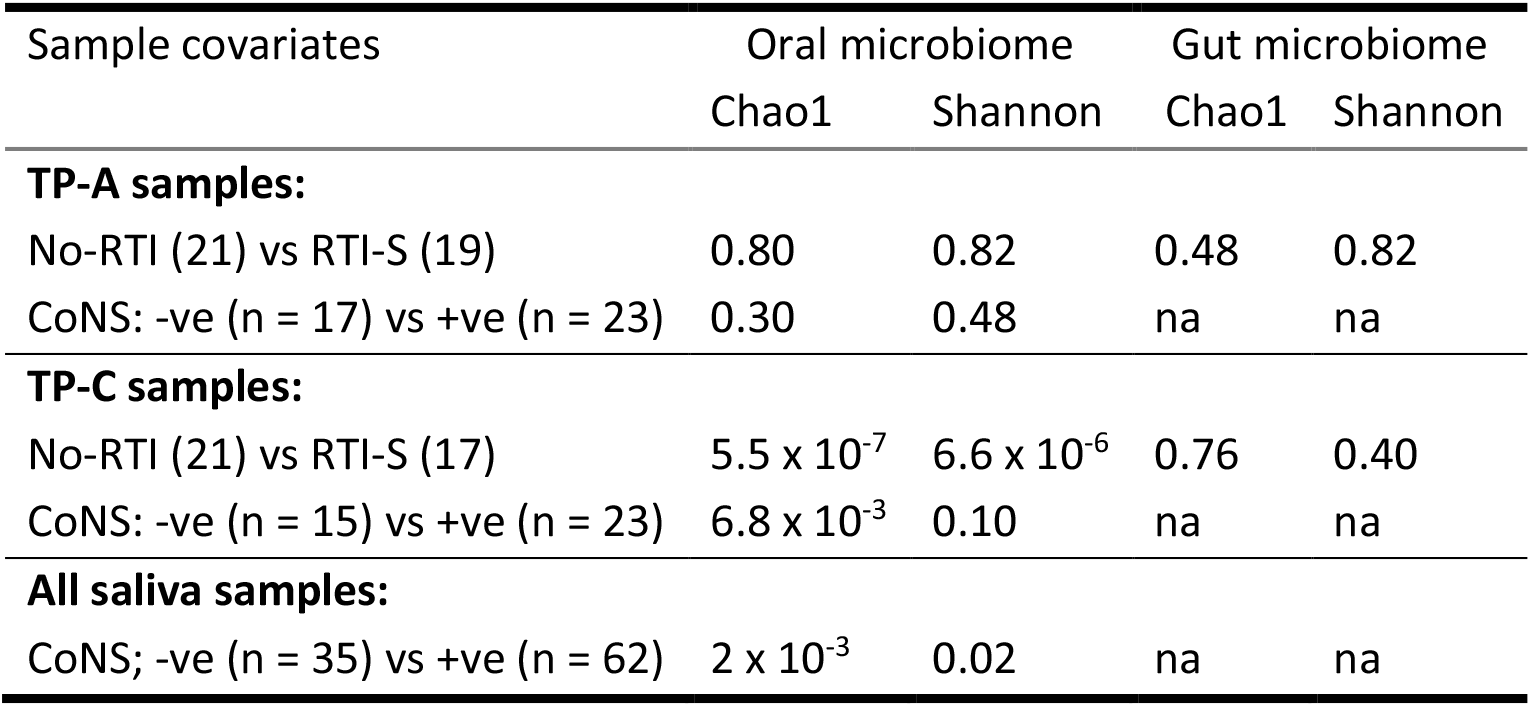
Oral and gut microbiome diversity in all samples and samples collected at time points A and C. Microbial richness was assessed using the Shannon diversity index and Chao1 measures, *p* values. Abbreviations: not applicable (na), RTI symptoms (RTI-S) and no RTI symptoms (no-RTI), coagulase-negative staphylococci (CoNS), time point (TP).

| Sample covariates | Oral microbiome |  | Gut microbiome |  |
| --- | --- | --- | --- | --- |
|  | Chao1 | Shannon | Chao1 | Shannon |
| **TP-A samples:** |  |  |  |  |
| No-RTI (21) vs RTI-S (19) | 0.80 | 0.82 | 0.48 | 0.82 |
| CoNS: -ve (n = 17) vs +ve (n = 23) | 0.30 | 0.48 | na | na |
| **TP-C samples:** |  |  |  |  |
| No-RTI (21) vs RTI-S (17) | $5.5 \times 10^{-7}$ | $6.6 \times 10^{-6}$ | 0.76 | 0.40 |
| CoNS: -ve (n = 15) vs +ve (n = 23) | $6.8 \times 10^{-3}$ | 0.10 | na | na |
| **All saliva samples:** |  |  |  |  |
| CoNS; -ve (n = 35) vs +ve (n = 62) | $2 \times 10^{-3}$ | 0.02 | na | na |

**Figure 4.**
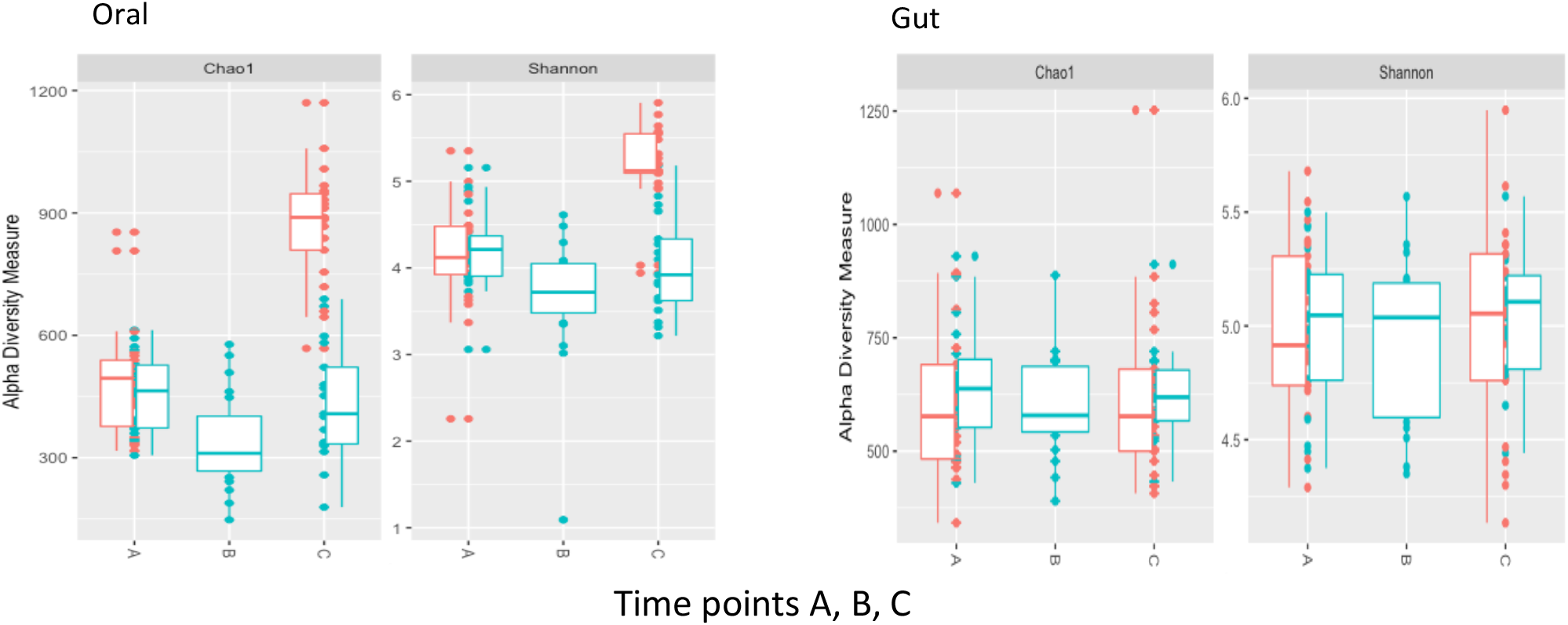
Alpha diversity in oral and gut microbiomes at time points A (baseline), B (during-RTI), C (post-RTI/end of study) in participants with RTI symptoms (RTI-S [blue]) and no RTI symptoms (no-RTI [coral]). Microbial richness was assessed by the Chao1 and Shannon diversity index measures.

Beta diversity, Bray-Curtis PCoA plots revealed no visually apparent clusters in any microbiomes at any time points. PERMANOVA tests for the OM indicated significant differences between RTI-S vs no-RTI participants (*p* = 0.001), CoNS positive vs CoNS negative (*p* = 0.002) whereas in the GM there were no significant differences between no-RTI vs RTI-S participants (*p* = 0.139).

#### Microbiome biomarkers

Microbial biomarkers were identified using LEfSe analysis based on the Kruskal-Wallis test and LDA (log10) for samples collected at the start of the study (TP-A) (Figure 5). The OM of RTI-S vs no-RTI participants showed an increased abundance of *Streptococcus sobrinus* and *Megamonas* and decreased abundance of Synergistetes, Verrucomicrobia, Dethiosulfovibrio and *Lactobacillus salivarius*. The GM of RTI-S vs no-RTI participants showed an increased abundance of *Rikenellaceae, Veillonella, Enhydobacteria, Eggerthella* and *Xanthomonsdales* and decreased abundance of *Desulfobulbus* and *Coprobacillus*.

**Figure 5.**
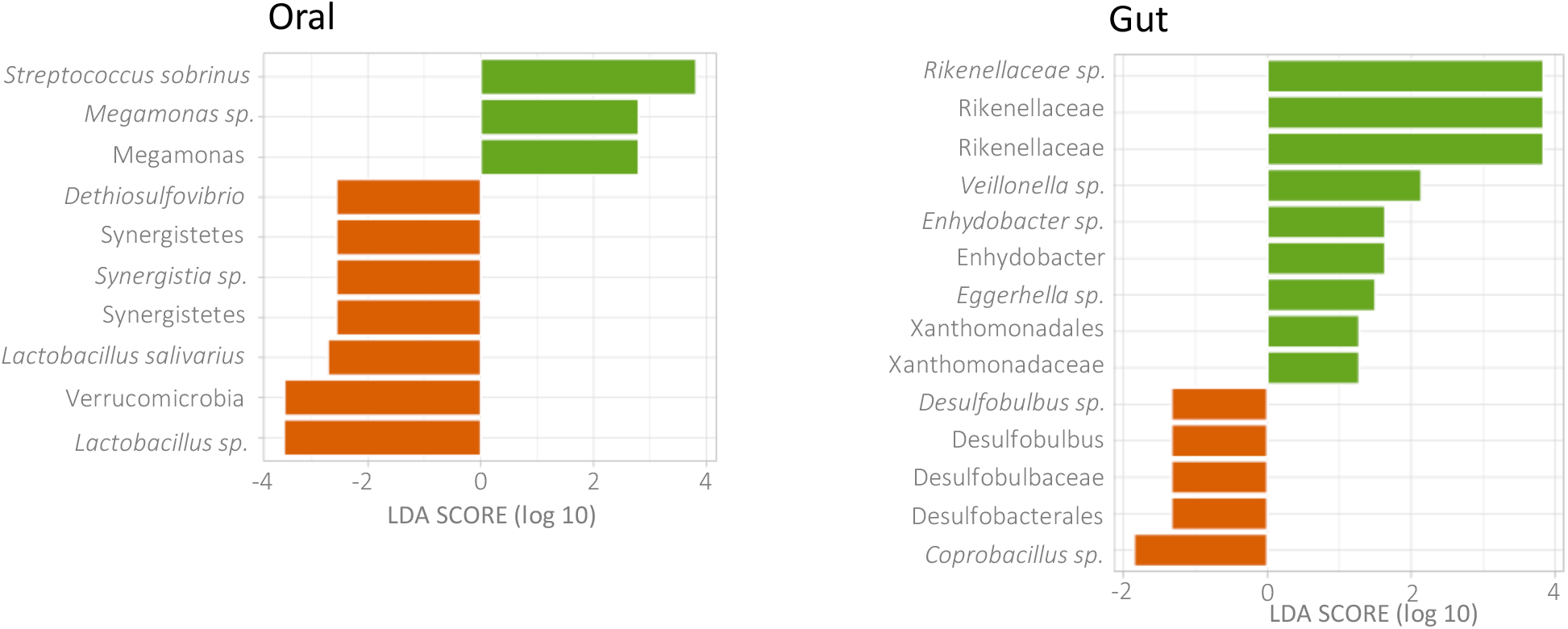
Biomarkers in oral and gut microbiomes from participants with RTI symptoms (RTI-S) compared those with no symptoms (no-RTI) at time point A generated from LEfSe generated LDA scores (log 10).

Taxa that best differentiate the RTI-S and no-RTI participant groups collected at the end of the study (TP-C) can be seen in supplementary Figure S2. The OM of RTI-S vs no-RTI participants showed an increased abundance of *Actinobacteria, Streptococcus, Lactobacillus* and decreased abundance of Ruminococcaceae and *Clostridia*. The GM of RTI-S vs no-RTI participants showed an increased abundance of *Ruminococcus torques, Catenibacterium* and *Clostridia* and a decreased abundance of *Eubacterium biforme, Erysipelotrichales* and *Coprobacillus*.

## Discussion

Our findings demonstrate the feasibility of conducting a longitudinal cohort microbiome study recruiting healthy adult participants from the community. We collected biological samples (stool and saliva samples proxy for gut and oral microbiomes [GM, OM]) for microbiome analysis, self-reported RTI symptoms and socio-demographic data. Microbiome biomarkers for susceptibility to RTIs were found by comparing the microbiomes of participants who developed RTI symptoms (RTI-S) to those who remained healthy (no-RTI) prior to RTI acquisition. Longitudinal microbiome tracking allowed a holistic approach furthering efforts to understand microbial variation changes during a RTI episode.

RTIs might arise from acquisition of an RTI pathogen, or from outgrowth of a pathobiont already present in the host. Understanding the role of the microbiome in these processes is an area of intense interest. Our findings identify that there were differences between the microbial composition of the microbiomes of those participants who acquired an RTI compared to those who did not, prior to the onset of RTI symptoms, but whether these went on to influence pathogen acquisition or pathobiont proliferation is unknown.

Strikingly, the carriage of CoNS was significantly greater in participants that acquired RTI compared to those who remained healthy. CoNS are opportunistic pathogens that colonize the skin and mucous membranes of the oral cavity in healthy individuals [23]. CoNS have been implicated as a significant etiological factor of RTIs including laryngological infections and viral infections such as chronic rhinosinusitis (CRS) (viral inflammation of the mucous membranes of the rhino-sinus) and pharyngitis (viral throat infection) yet disease mechanisms remain largely unknown [24] [25-27]. Although, there was no evidence to suggest CoNS caused the RTIs in participants, we found an association between CoNS with a less diverse microbiome and with participants who went on to acquire an RTI. The presence of CoNS as a potential biomarker of RTI susceptibility deserves further investigation.

We speculate that RTI symptoms were due to viral influenza and bacterial *H. influenzae*. Influenza (subtype A) and *H. influenzae* were common among saliva samples collected during an RTI, whereas *S. pneumoniae* and *M. catarrhalis* were of low prevalence. Seasonal community-acquired influenza-like illness (ILI) are common in adults [28] and influenza infection is often associated with synergistic secondary bacterial co-infection by *H. influenzae* [29]. Here, self-reported symptoms and RT-PCR results could have been strengthened by retrospective GP records. In addition, sample collection overlapped the UK pandemic, yet we found the SARS-CoV-2 virus wasn’t present in this cohort at this time.

Here, microbiome biomarkers of susceptibility to RTI in otherwise healthy adults were determined by linear discriminant analysis (LDA) effect size (LEfSe) to find differentially abundant taxa. In the OM of RTI-S participants a high abundance of *S. sobrinus* and Megamonas was found. *S. sobrinus* is a pathobiont that is strongly linked to the development of dental caries and oral disease [30]. In low abundance were Synergistetes, Lactobacillus (including *L. salivarius*) and Verrucomicrobia. Thus, in RTI-S participants beneficial bacteria such as *L. salivarius* with probiotic and antimicrobial properties are less dominant, perhaps contributing to the growth of pathobionts. A strong association between cross-niche network structure, key species, and subsequent susceptibility to RTI acquisition was found in infant nasopharyngeal, OM and GM [31]. Although early life microbiomes are immature in infancy and stable patterns may not be apparent until around 3-4 years of age [32], these studies are encouraging as they highlight the potential for the microbiota to influence RTI acquisition.

The GM was substantially more diverse than the oral cavity, as expected. In RTI-S participants, compared to no-RTI participants, there was a high abundance of Rikenellaceae, *Veillonella* and *Enhydrobacter* and decrease in the levels of *Coprobacillus. Veillonella* is associated with the GM of patients with COVID-19 compared to healthy controls [33]. In addition, this genus has been identified as a key microbe in colorectal associated-cancer and a putative gut biomarker for Cystic fibrosis [34]. It has been found that *Coprobacillus* was enriched in patients with COVID-19 [35] whereas our study participants did not have COVID-19. In older participants (aged 60 years) with ILI the gut biomarker *Ruminococcus torques* has been positively associated with infection risk [36]. In our study *R. torques* was enriched in RTI-S participants compared to no-RTI participants at the end of the study (TP-C). Our results suggest the gut niche may yield putative biomarkers of RTI acquisition which might subsequently aid in the rapid identification of those most vulnerable to infection in the community.

In the saliva OM two ‘stomatotypes’ exist, with stomatotype-1 characterised by abundant *Neisseria* and *Haemophilus* while stomatotype-2 is associated with a high level of *Prevotella* and *Veillonella* [37]. At the start of the study (TP-A) in healthy participants the core OM is indicative of stomatotype-1 but shifts to stomatotype-2 at the end of the study (TP-C) including an increase in pathobiont *P. melaninogenica*. An increased relative abundance of *P. melaninogenica* in the nasopharynx of children has been associated with severe influenza [38]. Core signatures can be useful for predicting oral health and possibly RTI [39]. It’s not clear why the OM of healthy participants undergoes a stomatotype shift, but perhaps the month when the samples were collected may have played a role (TP-A and TP-C sample collection was October and March respectively).

Here, we sought to test whether urban adult cohorts could be recruited to participate in microbiome studies, in order to investigate microbiome-RTI links for community acquired RTI. While we conclude that we have proved the feasibility of this approach, our study has limitations. The national lockdown in UK in response to the COVID-19 pandemic in March 2020 caused premature closure of this study, limiting the recruitment of participants and postal disruption resulted in several samples lost in transit. Furthermore, we were not able to collect health-related risk factors from medical notes. This study relied on self-reporting of RTI symptoms and severity, so access to medical history and current health records would have strengthened designation of RTI severity, identified prescribed medications, and enabled diagnostic confirmation of causative pathogen. However, this was not possible under lockdown restrictions. While our participant questionnaire was used to assess a broad range of microbiome covariables including dietary and lifestyle (alcohol intake, physical activity) factors, we observed microbiome diversity between the months of sample collection. Seasonal variation of the GM has been well documented and thus this factor must be taken into consideration in further seasonal RTI microbiome studies as a possible confounder [40]. We considered network microbiome analysis, however due to low participant numbers we predict this approach will lack statistical power when comparing network clusters across niches. We used RT-PCR to assess pathogen carriage as a microbiome covariable however microbe load was not considered as a covariate in this analysis. Finally, although we found individual response rates were low, once participants were recruited into the study retention rates tended to be high as seen in other RTI feasibility studies [41].

In conclusion, this feasibility study has shown that it is possible to recruit participants from the community, for them to collect stool and saliva samples and self-report RTI symptoms plus provide socio-demographic information for the duration of the 6-month study. Biomarker analysis of the oral and gut microbiomes before the onset of a RTI revealed compositional differences in those who acquired an RTI, where oral microbes were CoNS and *S. sobrinus* and gut bacteria of the genus *Veillonella* were distinguishing factors. This study contributes to the emerging picture that microbial biomarkers of susceptibility to RTI are identifiable, offering new insights into preventative measures.

## Supporting information

All supplementary information

## Data Availability

All data produced in the present study are available upon reasonable request to the corresponding author.

## References

1. Disease, G.B.D., I. Injury, and C. Prevalence, Global, regional, and national incidence, prevalence, and years lived with disability for 354 diseases and injuries for 195 countries and territories, 1990-2017: a systematic analysis for the Global Burden of Disease Study 2017. Lancet, 2018. 392(10159): p. 1789–1858.

2. Foundation, B.L. Economic Burden of Lung Disease. 2021; Available from: https://www.blf.org.uk/policy/economic-burden.

3. Confederation, N.H.S. Continuing costs of COVID-19. 2021; Available from: https://www.nhsconfed.org/publications/reckoning-continuing-cost-covid-19.

4. Enaud, R., et al., The Gut-Lung Axis in Health and Respiratory Diseases: A Place for Inter-Organ and Inter-Kingdom Crosstalks. Front Cell Infect Microbiol, 2020. 10: p. 9.

5. Chunxi, L., et al., The Gut Microbiota and Respiratory Diseases: New Evidence. J Immunol Res, 2020. 2020: p. 2340670.

6. Sarkar, A., et al., The gut microbiome as a biomarker of differential susceptibility to SARS-CoV-2. Trends Mol Med, 2021.

7. Caporaso, J.G., et al., QIIME allows analysis of high-throughput community sequencing data. Nat Methods, 2010. 7(5): p. 335–6.

8. Callahan, B.J., et al., DADA2: High-resolution sample inference from Illumina amplicon data. Nat Methods, 2016. 13(7): p. 581–3.

9. Katoh, K., et al., MAFFT: a novel method for rapid multiple sequence alignment based on fast Fourier transform. Nucleic Acids Res, 2002. 30(14): p. 3059–66.

10. Price, M.N., P.S. Dehal, and A.P. Arkin, FastTree 2--approximately maximum-likelihood trees for large alignments. PLoS One, 2010. 5(3): p. e9490.

11. Bokulich, N.A., et al., Optimizing taxonomic classification of marker-gene amplicon sequences with QIIME 2’s q2-feature-classifier plugin. Microbiome, 2018. 6(1): p. 90.

12. McDonald, D., et al., An improved Greengenes taxonomy with explicit ranks for ecological and evolutionary analyses of bacteria and archaea. ISME J, 2012. 6(3): p. 610–8.

13. Bisanz, J.E., qiime2R: Importing QIIME2 artifacts and associated data into R sessions. 2018.

14. Team, R.C., R: a language and environment for statistical computing. 2014: R Foundation for Statistical Computing.

15. Wickham, H., ggplot2: Elegant Graphics for Data Analysis. 2016, New York: Springer-Verlag.

16. McMurdie, P.J. and S. Holmes, phyloseq: an R package for reproducible interactive analysis and graphics of microbiome census data. PLoS One, 2013. 8(4): p. e61217.

17. Callahan, B.J., et al., Bioconductor Workflow for Microbiome Data Analysis: from raw reads to community analyses. F1000Res, 2016. 5: p. 1492.

18. Davis, N.M., et al., Simple statistical identification and removal of contaminant sequences in marker-gene and metagenomics data. Microbiome, 2018. 6(1): p. 226.

19. Leo Lahti, S.S.e.a. Tools for microbiome analysis in R. Version 1.5.23. 2017; Available from: https://microbiome.github.io/tutorials/Core.html.

20. Segata, N., et al., Metagenomic biomarker discovery and explanation. Genome Biol, 2011. 12(6): p. R60.

21. Jones, N.K., et al., Evaluating the use of a 22-pathogen TaqMan array card for rapid diagnosis of respiratory pathogens in intensive care. J Med Microbiol, 2020. 69(7): p. 971–978.

22. Ordonez-Mena, J.M., et al., Relationship between microbiology of throat swab and clinical course among primary care patients with acute cough: a prospective cohort study. Fam Pract, 2020. 37(3): p. 332–339.

23. Heilmann, C., W. Ziebuhr, and K. Becker, Are coagulase-negative staphylococci virulent? Clin Microbiol Infect, 2019. 25(9): p. 1071–1080.

24. Michalik, M., et al., Coagulase-negative staphylococci (CoNS) as a significant etiological factor of laryngological infections: a review. Ann Clin Microbiol Antimicrob, 2020. 19(1): p. 26.

25. Becker, K., C. Heilmann, and G. Peters, Coagulase-negative staphylococci. Clin Microbiol Rev, 2014. 27(4): p. 870–926.

26. Dlugaszewska, J., et al., The pathophysiological role of bacterial biofilms in chronic sinusitis. Eur Arch Otorhinolaryngol, 2016. 273(8): p. 1989–94.

27. Gudima, I.A., et al., [Viral-bacterial-fungal associations in chronic tonsillitis in children]. Zh Mikrobiol Epidemiol Immunobiol, 2001(5): p. 16–9.

28. Tellioglu, E., G. Balci, and A. Mertoglu, Duration of Stay of Patients with Community-Acquired Pneumonia in Influenza Season. Turk Thorac J, 2018. 19(4): p. 182–186.

29. Morris, D.E., D.W. Cleary, and S.C. Clarke, Secondary Bacterial Infections Associated with Influenza Pandemics. Front Microbiol, 2017. 8: p. 1041.

30. Li, J.W., R.M. Wyllie, and P.A. Jensen, A Novel Competence Pathway in the Oral Pathogen Streptococcus sobrinus. J Dent Res, 2021. 100(5): p. 542–548.

31. Reyman, M., et al., Microbial community networks across body sites are associated with susceptibility to respiratory infections in infants. Commun Biol, 2021. 4(1): p. 1233.

32. Kumbhare, S.V., et al., Factors influencing the gut microbiome in children: from infancy to childhood. J Biosci, 2019. 44(2).

33. Gu, S., et al., Alterations of the Gut Microbiota in Patients With Coronavirus Disease 2019 or H1N1 Influenza. Clin Infect Dis, 2020. 71(10): p. 2669–2678.

34. Dayama, G., et al., Interactions between the gut microbiome and host gene regulation in cystic fibrosis. Genome Med, 2020. 12(1): p. 12.

35. Zuo, T., et al., Alterations in Gut Microbiota of Patients With COVID-19 During Time of Hospitalization. Gastroenterology, 2020.

36. Susana Fuentes, G.d.H., Nening M Nanlohy, Lucas Wijnands, José A Ferreira, Mioara A Nicolaie, Jeroen L A Pennings, Ronald Jacobi, and J.v.B. Jelle de Wit, Debbie van Baarle, Associations of faecal microbiota with influenza-like illness in participants aged 60 years or older: an observational study. Lancet Healthy Longevity, 2021. 2: p. 13–23.

37. Willis, J.R., et al., Citizen science charts two major “stomatotypes” in the oral microbiome of adolescents and reveals links with habits and drinking water composition. Microbiome, 2018. 6(1): p. 218.

38. Langevin, S., et al., Early nasopharyngeal microbial signature associated with severe influenza in children: a retrospective pilot study. J Gen Virol, 2017. 98(10): p. 2425–2437.

39. Lenartova, M., et al., The Oral Microbiome in Periodontal Health. Front Cell Infect Microbiol, 2021. 11: p. 629723.

40. Koliada, A., et al., Seasonal variation in gut microbiota composition: cross-sectional evidence from Ukrainian population. BMC Microbiol, 2020. 20(1): p. 100.

41. Anderson, E.C., et al., Population-based paediatric respiratory infection surveillance: a prospective inception feasibility cohort study. Pilot Feasibility Stud, 2018. 4: p. 182.

