## Supplementary material for "Oral and gut microbiome biomarkers of susceptibility to respiratory tract infection in adults: a longitudinal cohort feasibility study": All supplementary information

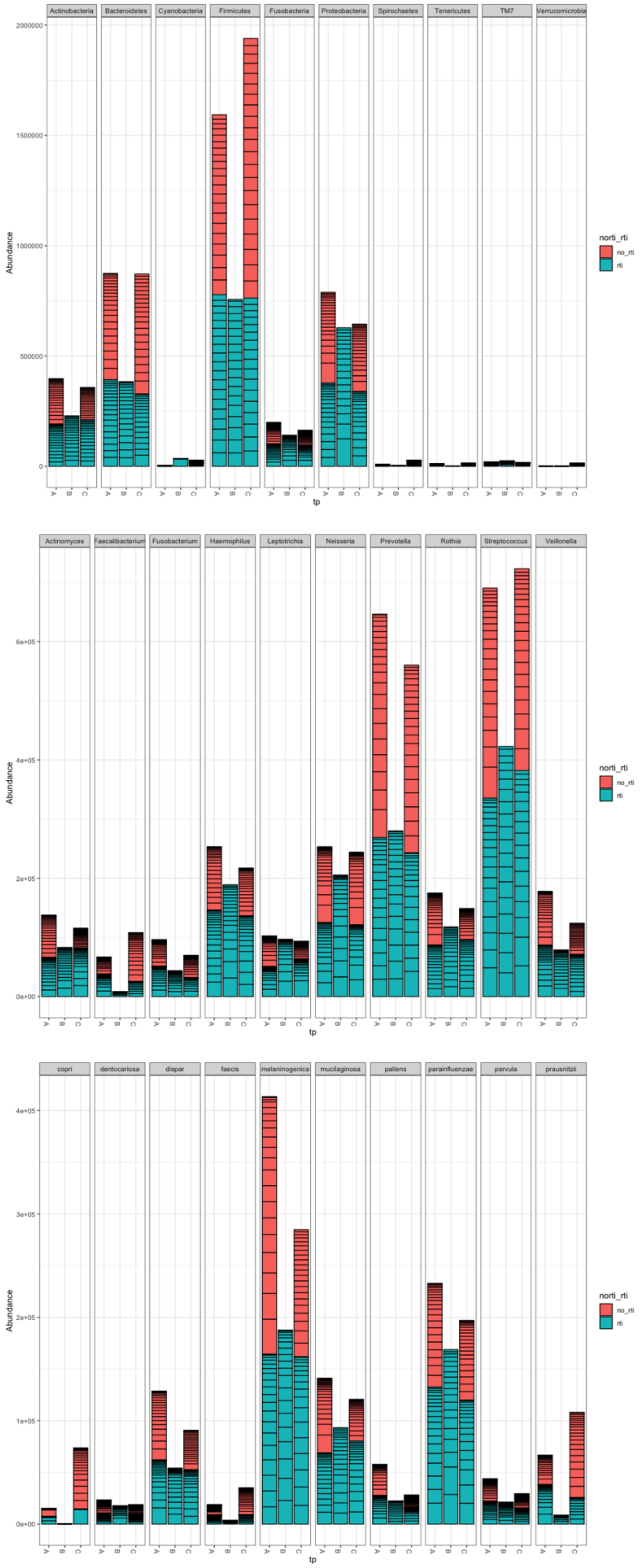


Supplementary Figure S1A. Oral microbiome.

The top 10 phyla (top), genus (middle) and species (bottom). On the y axis the data is displayed at each time point A, B, C, showing the relative proportion of no-RTI and RTI-S samples.

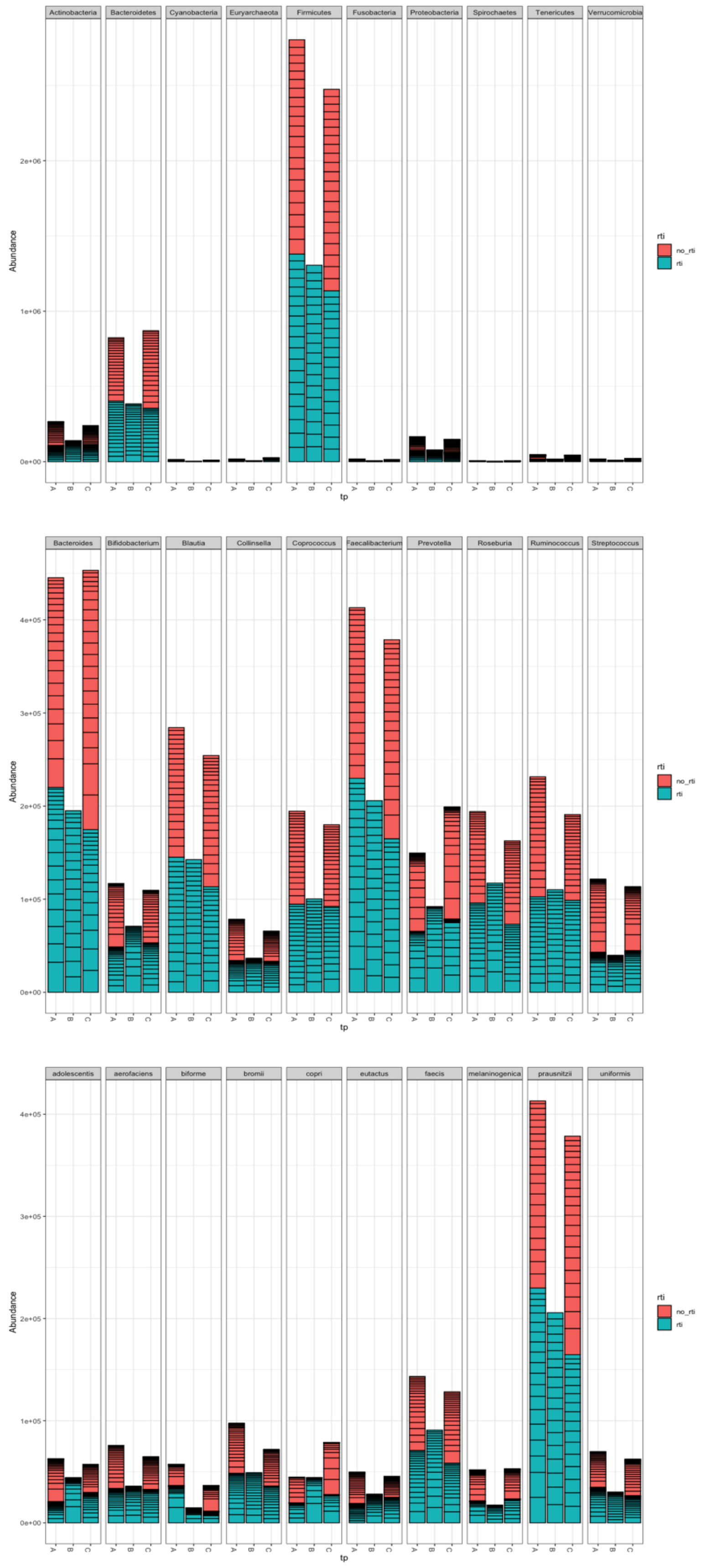


Supplementary Figure S1B. Gut microbiome.

The top 10 phyla (top), genus (middle) and species (bottom). On the y axis the data is displayed at each time point A, B, C, showing the proportion of no-RTI and RTI-S samples.

| Covariates of interest | Oral | | Gut | |
|---|---|---|---|---|
| | Chao1, *p* | Shannon, *p* | Chao1, *p* | Shannon, *p* |
| **All samples:** | | | | |
| December (n = 15), January (n = 49) | 0.72 | 0.15 | $7 \times 10^{-4}$ | $8 \times 10^{-6}$ |
| January (n = 49), February (n = 29) | 0.93 | 0.75 | $2 \times 10^{-4}$ | $5.4 \times 10^{-5}$ |
| Employed (n = 72), retired (n = 20)* | 0.71 | 0.59 | 0.03 | 0.01 |
| Pets no (n = 51), yes (n = 43)* | 0.47 | 0.40 | $7.9 \times 10^{-4}$ | $7.2 \times 10^{-3}$ |
| **In saliva only:** | | | | |
| CoNS; -ve (n = 35), +ve (n = 62) | $2 \times 10^{-3}$ | 0.02 | na | na |
| *H. influenzae*: -ve (n = 74), +ve (n = 23) | 0.40 | 0.74 | na | na |
| *S. pneumoniae*: -ve (n = 79), +ve (n = 18) | 0.73 | 0.88 | na | na |
| *M. catarrhalis*: -ve (n = 86), +ve (n = 11) | 0.14 | 0.19 | na | na |

Supplementary Table S1. Alpha diversity of covariates of interest in stool and saliva from participants with RTI symptoms and those that remained healthy.
*some data was lost due to pandemic distruptions.

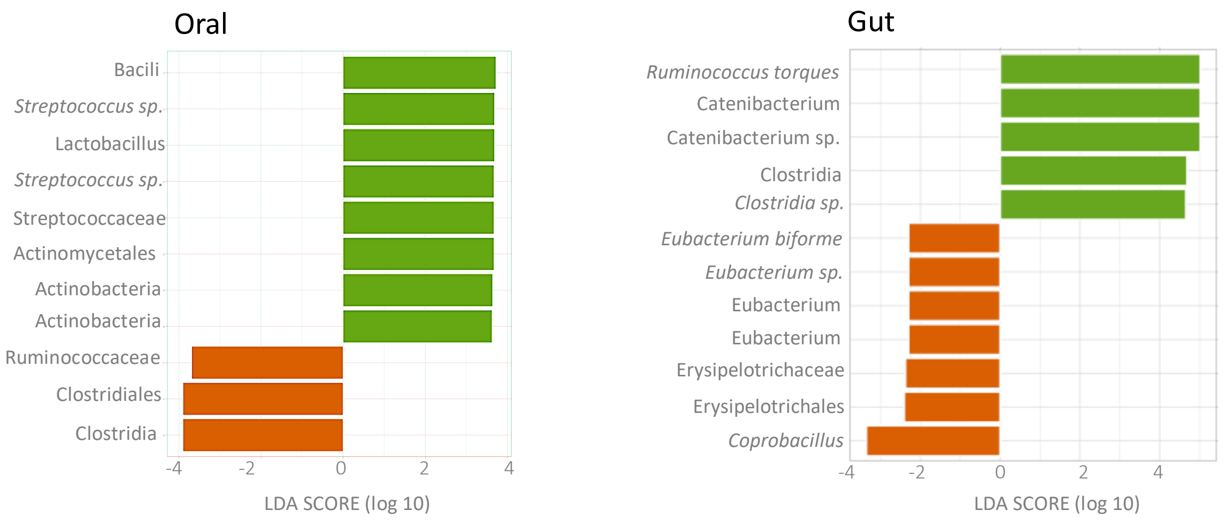


Supplementary Figure S2. Biomarkers in oral and gut microbiomes from participants with RTI symptoms (RTI-S) compared those with no symptoms (no-RTI) at time point C generated from LEfSe generated LDA scores (log 10).
